# Determinants of healthcare employees’ preference to continue teleworking after the COVID-19 pandemic: a cross-sectional study using hierarchical regression

**DOI:** 10.1101/2022.07.18.22277694

**Authors:** Andrea Marie Jones, Jonathan Fan, Leah Thomas-Olson, Wei Zhang, Christopher B McLeod

**Affiliations:** Partnership for Work Health and Safety, School of Population and Public Health, University of British Columbia, Vancouver, British Columbia, Canada; Fraser Health Authority, Surrey, British Columbia, Canada; Centre for Health Evaluation and Outcome Sciences, St. Paul’s Hospital, Vancouver, British Columbia, Canada; Institute for Work and Health, Toronto, Ontario, Canada

**Keywords:** healthcare workforce, work from home, telework, work arrangement, remote work

## Abstract

Employees’ post-pandemic telework preference is an important consideration for navigating post-pandemic work arrangements and can inform organizational planning and workforce management. A cross-sectional survey of employees (n=400, participation rate =36.4%) of a regional health authority who teleworked during the COVID-19 pandemic was conducted. The most common post-pandemic telework preference was all the time (52%) followed by over half but not all the time (32%) and less than half the time or not at all (16%). Using hierarchical multinomial logistic regression models and less than half the time or not at all as the reference outcome, being a provider of direct patient care and productivity while teleworking were strong determinants of post-pandemic telework preference while two or more weekly teleconference hours, work-life balance and having one or more people over five years of age in the home while teleworking were moderate determinants.

## Introduction

In April 2020, a rapid global shift from traditional on-site work to less traditional telework occurred in response to the COVID-19 pandemic in many sectors, including healthcare. 1 During the first year of the pandemic, approximately 17% of Canadian healthcare employees performed most or all of their work hours via telework. 2 At the current time, approximately two years following the pandemic onset, many physical distancing and stay at home orders and recommendations have been removed or made less stringent, creating increased opportunity for return to on-site work. Healthcare employee preference to continue teleworking post-pandemic and the determinants of this preference, are important considerations for navigating establishment of future work arrangements as well as broader healthcare organizational planning and workforce management.

A survey of corporate and non-clinical employees of a large local health district in Sydney Australia who teleworked as a response to the pandemic, found that 90% would take the opportunity to work from home again if offered. 3 Multi-sector surveys also indicate that many employees prefer to continue teleworking post-pandemic. A survey of Canadian employees who newly started teleworking during the pandemic found that 91% preferred to telework for at least some work hours post-pandemic. 4 Similar surveys found that 54% to 78% of American employees whose job can be performed via telework 5,6, and 72% to 82% of Hong Kong employees 7,8 and 39% of Vietnamese men and 63% of Vietnamese women who teleworked during the pandemic, would like to continue teleworking post-pandemic. 9 A secondary finding from these studies is that flexible hybrid work arrangements with telework and on-site components were more commonly preferred for the post-pandemic period than exclusive telework arrangements. 5-11 An exception is an American survey where hybrid and exclusive telework arrangements were equally as common. 5

Two studies on the determinants of employees’ post-pandemic telework preference were identified. Among Canadian employees, higher productivity while teleworking was a strong determinant of preferring to telework for most or all work hours post-pandemic while being a teacher was a strong determinant of preferring to work most or all hours on-site. 11 Among Vietnamese employees, valuing telework as a solution to air quality issues and having workaholic characteristics such as working late or overtime were positively associated with a preference to telework post-pandemic for both men and women while finding pleasure in elements of on-site work such as in-person collaboration or communication with colleagues was negatively associated. 9 Gender specific determinants were also found among Vietnamese employees. Among men only, having a graduate level education and a positive perception of telework were associated with a preference to telework post-pandemic; and among women only, middle age employees were less likely to prefer to telework post-pandemic than their younger counterparts and having two or more children in the home was associated with a preference for a hybrid work arrangement post-pandemic. 9

Literature from prior to the pandemic can help us to identify potential determinants of employees’ post-pandemic telework preference, although determinants may vary with time and context. A study of University employees in Malaysia conducted shortly prior to the pandemic found that both personal and job-level factors were important determinants of preferring to telework. 12 At the personal level, having a higher number of young children in the home and a longer morning commute, while at the job level less frequent use of face-to-face communication and fax machines and more frequent use of email and cell phones were important determinants. 12 Potential advantages and disadvantages of telework have been a focus of the pre-pandemic literature and these may influence telework preference. Identified advantages include better balance of work and home life, easier completion of domestic chores, increased work flexibility and autonomy, reduced commuting time and expenses, increased productivity, fewer distractions or interruptions, higher morale and job satisfaction, the ability to work while sick, decreased need for disability related accommodation, and avoidance of office politics. 13,14 Identified disadvantages include blurring of boundaries between work and home time, overwork or longer working hours, less sick leave, social isolation, lack of support, inadequate equipment, lack of career progression or promotions, and resentment from colleagues. 13,14 Each of these advantages and disadvantages will likely persist post-pandemic but due to changing telework norms and practices, the extent of their influence on telework preference in the post-pandemic period may vary from pre-pandemic.

The current study expands knowledge of employee telework preference and determinants to the post-pandemic period with a focus on the healthcare sector. Preliminary multi-sectoral evidence indicates that many employees will prefer to continue teleworking post-pandemic 5–11; as does a single Australian study of corporate and non-clinical healthcare employees. 3 Building further knowledge of post-pandemic telework preference and determinants in the healthcare sector is significant as healthcare is a major employment sector in Canada and around the world, representing approximately 13% of all employees in British Columbia. 15 The healthcare sector has traditionally had low telework capacity compared to other sectors. 16 Continuation of telework practices established during the pandemic in healthcare could represent a meaningful long-term shift in healthcare work practices and culture, including a shift towards virtualization of healthcare delivery. Lastly, healthcare employees have faced unprecedented challenges during the pandemic with pandemic-related occupational stressors contributing to poor general well-being and occupational burnout. 17,18 According to person-environment fit theory, compatibility between employee need and the demands or opportunities presented by the work environment contributes to the development of positive health and occupational outcomes. 19,20 Understanding healthcare employees’ preferred post-pandemic work arrangements might help healthcare organizations to maximize person-environment fit and subsequently to recruit and retain employees, as well as to offset other potential negative effects of occupational pandemic-related stressors.

The aims of this study were two-fold. Using employees of a large healthcare authority in British Columbia, Canada who teleworked at some point during the pandemic as the target population, the aims were to identify 1) the proportion of employees with a preference for a post-pandemic work arrangement that is: exclusively telework, mostly telework, mostly on-site, or entirely on-site and 2) sociodemographic and occupational characteristics and e-working conditions and practices associated with employees’ preference to continue teleworking after the pandemic.

## Methods

### Study sample

An observational cross-sectional study design was used. The target population included clinical and non-clinical employees of a large regional health authority in British Columbia, Canada who teleworked at some point during the COVID-19 pandemic. The organization includes nearly 40,000 employees, and is responsible for providing hospital care, community-based long-term care, home health, mental health and public health services to a large distributed geographic region. It was selected for the study in response to strong interest among the organization’s senior leadership to understand employees’ health needs while teleworking during the third pandemic wave, as well as employees’ telework preferences for the post-pandemic period.

Data for this study was collected using a voluntary survey distributed via Qualtrics 21 with nearly 100 items on demographics, occupation, e-working conditions, and health. Participants were recruited between May and June 2021, using internal newsletters and emails endorsed by senior leadership. Managers were encouraged to support participation and employees were able to complete the survey using time in lieu. All participants provided informed consent using a virtual consent form prior to the online survey. Ethical approval was provided by the UBC Behavioural Research Ethics Board (#H21-00434) and the Fraser Health Research Ethics Board (#FHREB 2021-030).

### Study Variables

#### Post-pandemic telework preference

For the outcome variable, participants were asked “Would you like to continue working from home or performing virtual work following the end of the COVID-19 pandemic?” with four possible response options: “No, not at all”, “Yes, but for less than half of the time”, “Yes, for more than half of the time”, and “Yes, all the time”. ‘Virtual work’ was included in the telework outcome to capture work that i) was performed at an off-site remote location other than the employee’s primary residence (e.g. a short-term vacation rental) and ii) adapted use of information technology to communicate with patients or colleagues in place of in person communication as a response to the pandemic, although this work may have been performed in a clinic or hospital and not at home. Due to a small number of respondents (n=5) who indicated “No, not at all”, these respondents were grouped with those who answered “Yes, but for less than half the time” for analysis.

#### Sociodemographic characteristics

Sociodemographic characteristics included gender identity, age group, number of children under 5 years of age, number of other people over 5 years of age in the home during work hours, and having a disability or long term health condition.

#### Occupational characteristics

Occupational characteristics included having direct reports, full time job status, providing direct patient care, prior work telework experience, and years worked at the organization. For gender identity, due to a small number of non-binary respondents (less than 5), non-binary and women respondents were grouped together to maintain anonymity. 22

#### E-working conditions

Psychosocial e-working conditions were measured using items from the validated e-work life scale 23 as well as a single item on time spent on teleconferencing. The E-work life scale is a 17-item scale with four subscales that measure theoretically important aspects of the psychosocial e-working experience: work-life balance (7 items), productivity (4 items), organizational trust (3 items) and flexibility (3 items) (see Table 1). Participants were instructed that for the purposes of the survey, the term ‘e-working’ as it appears in the e-work life scale items refers to both work performed from home or virtually, including virtual work performed in a clinic or hospital. Item responses were recorded using a 5-point Likert scale (from 1 = strongly agree to 5 – strongly disagree with an option of ‘N/A or no opinion’). The score for each subscale was calculated by taking the average of the scores from the corresponding items. If one response was ‘N/A or no opinion’ within a subscale, the mean score among the remaining items was used. We considered the subscale missing if there was more than one response of ‘N/A or no opinion’. Reverse scoring was used for 4 items in the work life balance subscale such that all higher scores consistently reflected a better e-work experience across all items. Time spent using video or teleconference technology was measured as the average numbers of weekly hours.

**Table 1.**
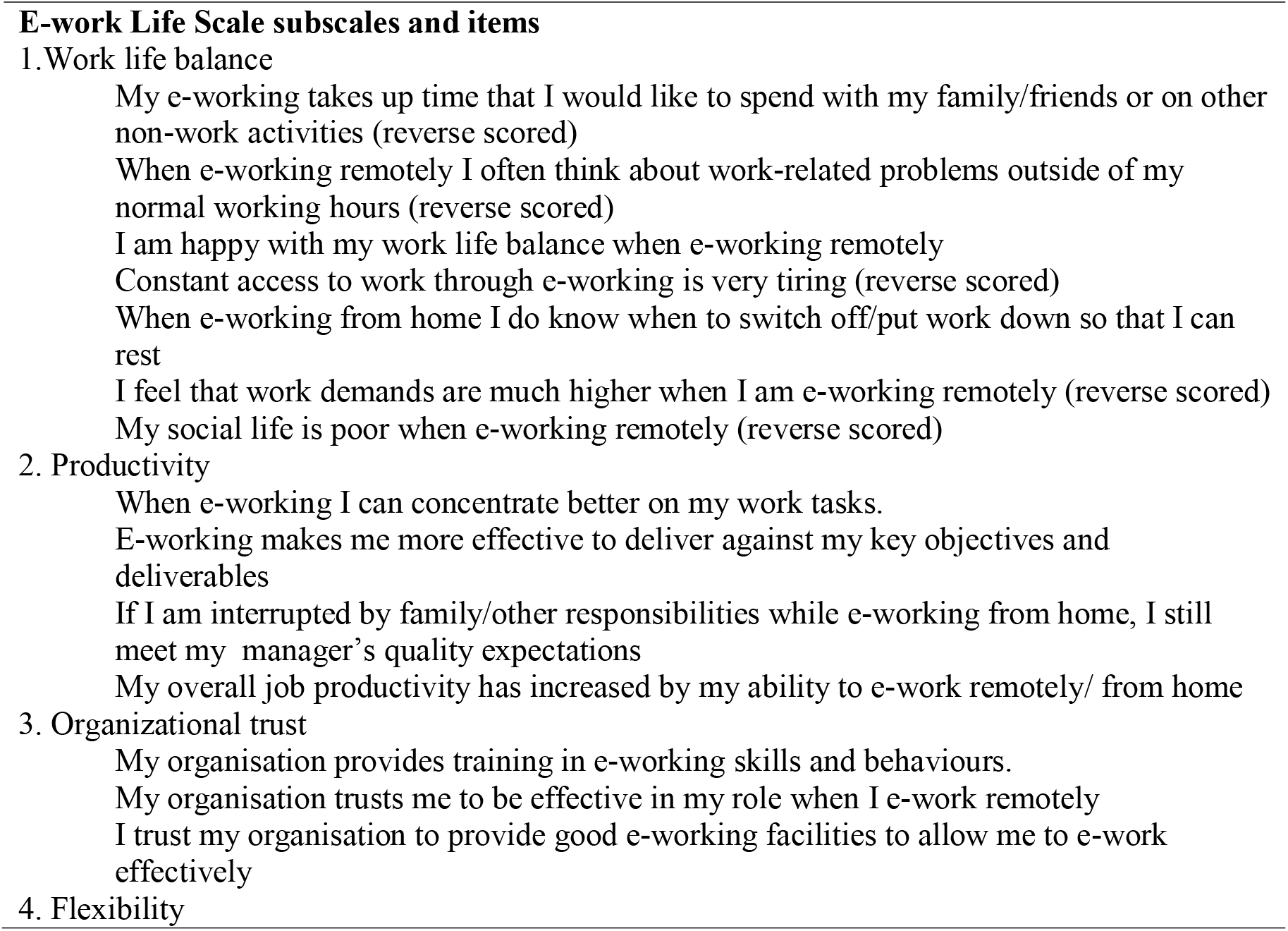
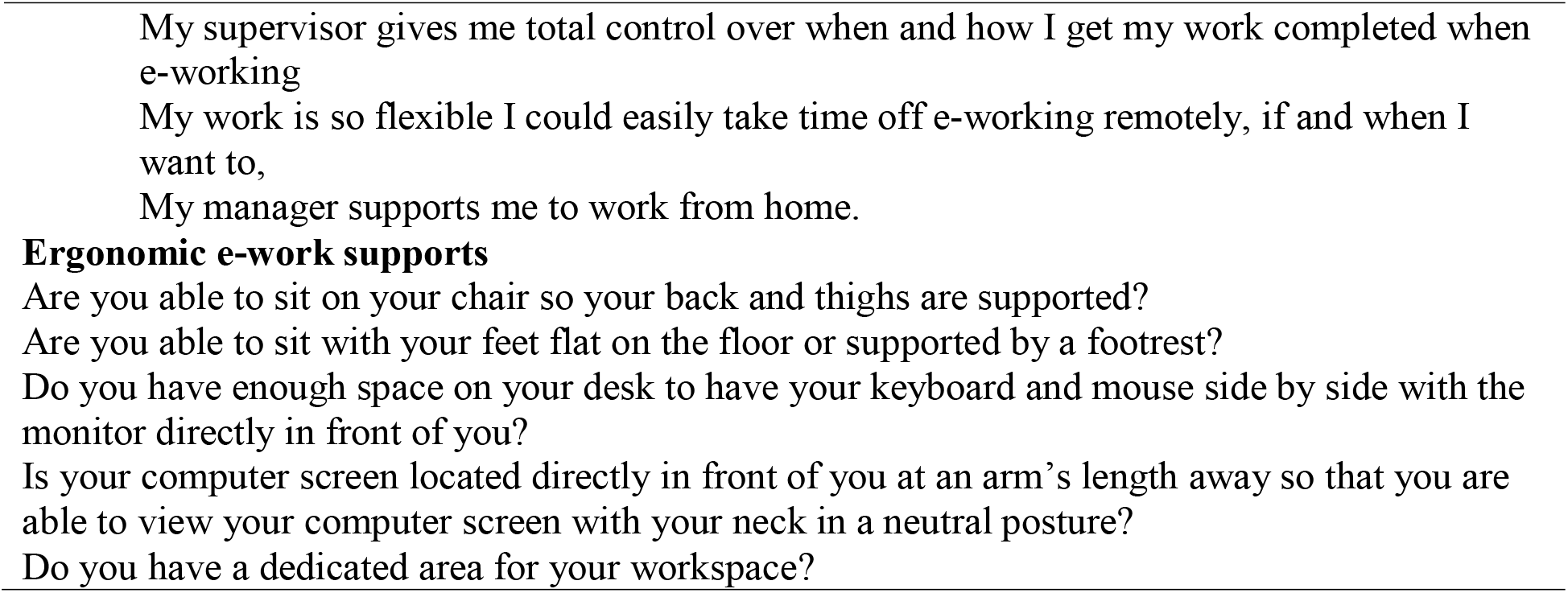
Description of e-work life scale subscales and items and ergonomic e-work items included on survey

Ergonomic e-work conditions were measured using five binary (yes/no) items adapted from the employer’s internal ergonomic assessment tool (see Table 1). A single binary ergonomic score was computed where participants were coded as having answered ‘yes’ to all 5 ergonomics items versus participants who answered ‘no’ to at least one ergonomic item.

#### Analyses

Frequencies and proportions were calculated for the study variables for the study sample overall and by telework preference. In line with recommendations for multiple exposure studies, we used a hierarchical multiple regression approach to identify determinants of telework preference as well as to account for their hierarchical nature. 24 Potential determinants were conceptually grouped into discrete blocks based on similarity, and then the blocks were conceptually organized based on proximity to the outcome. This resulted in three blocks ordered from most distal to most proximal to the outcome as follows: sociodemographic characteristics, occupational characteristics, and e-working conditions. Hierarchical approaches such as this are appropriate for models that contain both distal and proximal determinants, where the more distal determinants might influence not only the outcome directly, but also any other determinants that are relatively more proximal to the outcome. In such situations, addition of all determinants into one single adjusted model, can bias effect estimates for determinants that are more distal to the outcome than others. Hierarchical regression ensures that the estimates of the impacts of more distal determinants on the outcome are not biased by including more proximal determinants in the same model. It also ensures that the potential confounding effects of more distal determinants are accounted for in the estimates of the impacts of more proximal determinants on the outcome.

Ordinal logistic regression models with a 3 level outcome variable (less than half the time or not at all, more than half the time but not all the time, and all the time) were conducted but the proportional odds assumption was not met for several variables (data not shown). 25 Next, multinomial logistic regression models were conducted using “less than half the time or not at all” as the reference group (25). Unadjusted models for each independent variable and the outcome were conducted. For the adjusted models, a three-step hierarchical approach was used with personal-level sociodemographic and occupational variables added as the first and second blocks respectively, and e-working variables added as the third block. Odds ratios (OR) and 95% confidence intervals (CI) are reported for all multinomial logistic regression models. Odds radios below 0.5 or above 2.0 were considered moderate to strong. 26 Lastly, to examine if the addition of each successive block improved the goodness-of-fit in the model over and beyond the previous block(s), the Akaike information criterion (AIC) value was reported for each block adjusted model. 27

## Results

A total of 453 of an estimated 1100 employees who teleworked at some point during the pandemic answered the survey. Of these 53 (11.7%) were excluded due to missing data for one or more study variables. The final sample included 400 respondents for a 36.4% participation rate.

The final study sample was 83.5% women or non-binary people (Table 2). The most common age group was 46 to 35 years of age (28.8%), with slightly less respondents in the 35 years or less (26.0%) and 46 to 55 years of age (28.8%) groups, and even fewer in the 56 years of age or more group (14.5%). During working hours, less than a quarter (23.3%) of respondents had children under 5 years of age and slightly over half (53.0%) had at least one person over 5 years of age in the home. Only 13.5% of respondents identified as having a disability or long term health condition.

**Table 2.**
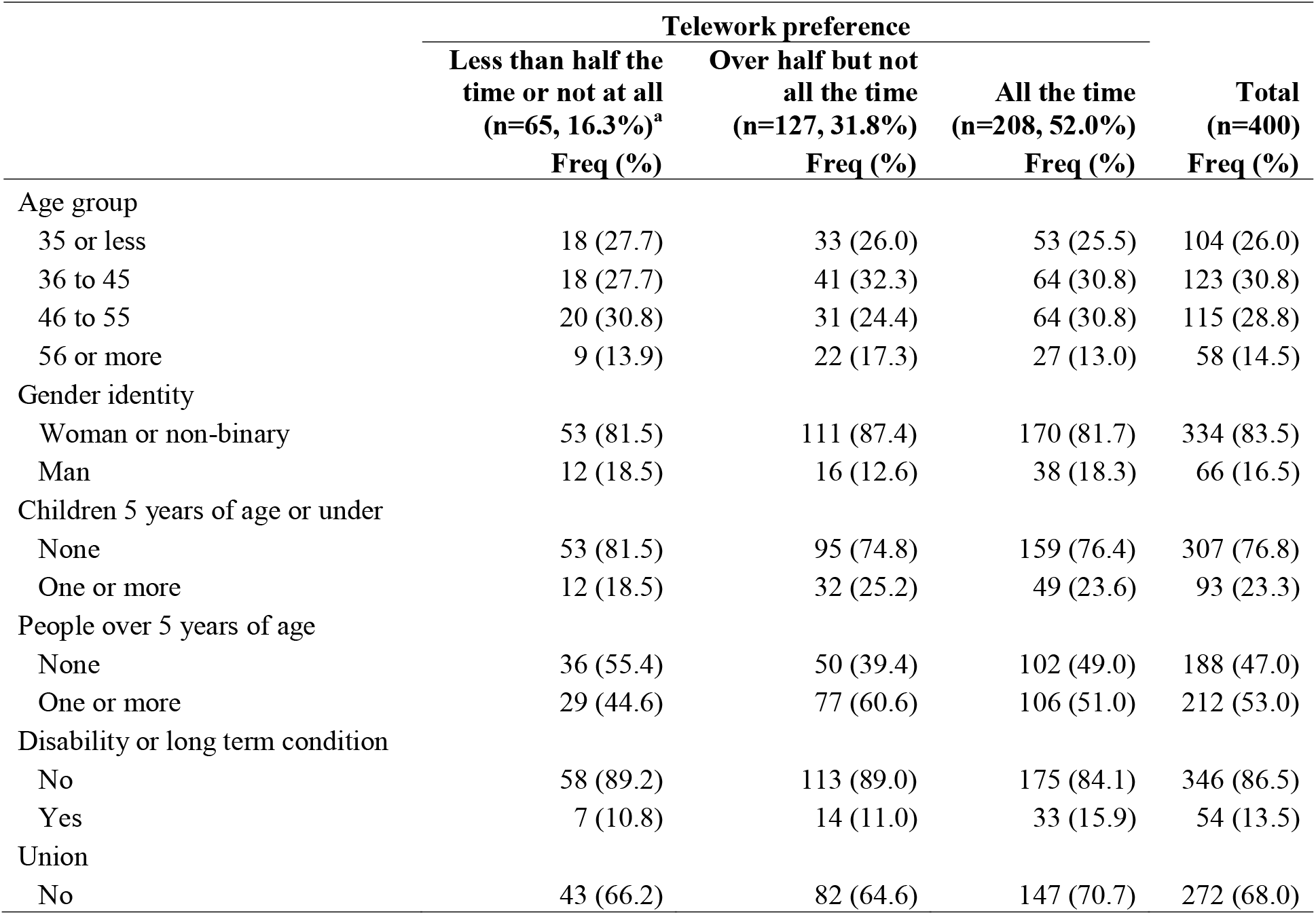
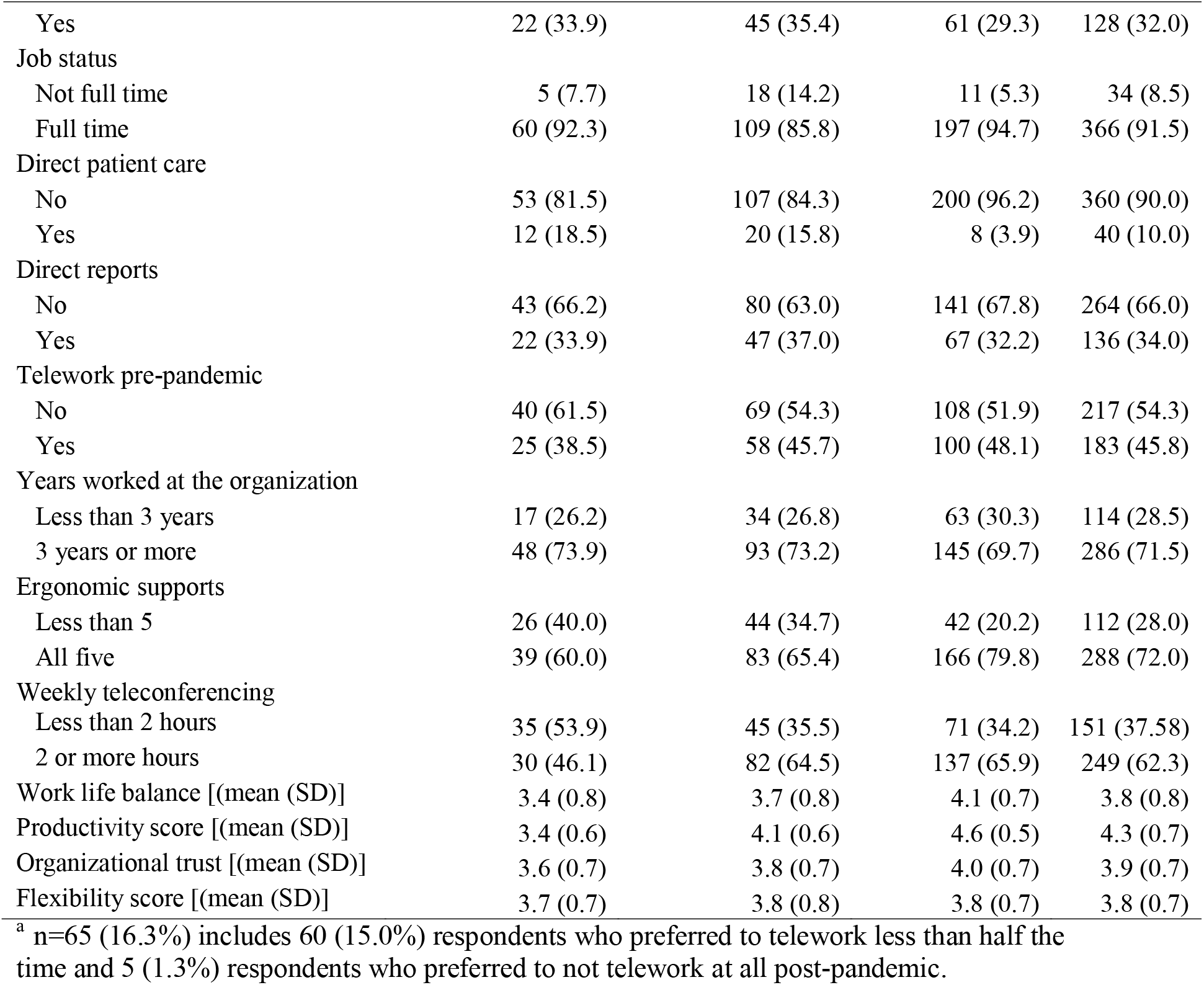
Characteristics of the analytic sample overall and by telework preference

|  | Telework preference |  |  | Total<br>(n=400)<br>Freq (%) |
| --- | --- | --- | --- | --- |
|  | Less than half the<br>time or not at all<br>(n=65, 16.3%)<sup>a</sup> | Over half but not<br>all the time<br>(n=127, 31.8%) | All the time<br>(n=208, 52.0%) |  |
|  | Freq (%) | Freq (%) | Freq (%) |  |
| Age group |  |  |  |  |
| 35 or less | 18 (27.7) | 33 (26.0) | 53 (25.5) | 104 (26.0) |
| 36 to 45 | 18 (27.7) | 41 (32.3) | 64 (30.8) | 123 (30.8) |
| 46 to 55 | 20 (30.8) | 31 (24.4) | 64 (30.8) | 115 (28.8) |
| 56 or more | 9 (13.9) | 22 (17.3) | 27 (13.0) | 58 (14.5) |
| Gender identity |  |  |  |  |
| Woman or non-binary | 53 (81.5) | 111 (87.4) | 170 (81.7) | 334 (83.5) |
| Man | 12 (18.5) | 16 (12.6) | 38 (18.3) | 66 (16.5) |
| Children 5 years of age or under |  |  |  |  |
| None | 53 (81.5) | 95 (74.8) | 159 (76.4) | 307 (76.8) |
| One or more | 12 (18.5) | 32 (25.2) | 49 (23.6) | 93 (23.3) |
| People over 5 years of age |  |  |  |  |
| None | 36 (55.4) | 50 (39.4) | 102 (49.0) | 188 (47.0) |
| One or more | 29 (44.6) | 77 (60.6) | 106 (51.0) | 212 (53.0) |
| Disability or long term condition |  |  |  |  |
| No | 58 (89.2) | 113 (89.0) | 175 (84.1) | 346 (86.5) |
| Yes | 7 (10.8) | 14 (11.0) | 33 (15.9) | 54 (13.5) |
| Union |  |  |  |  |
| No | 43 (66.2) | 82 (64.6) | 147 (70.7) | 272 (68.0) |
| Yes | 22 (33.9) | 45 (35.4) | 61 (29.3) | 128 (32.0) |
| Job status |  |  |  |  |
| Not full time | 5 (7.7) | 18 (14.2) | 11 (5.3) | 34 (8.5) |
| Full time | 60 (92.3) | 109 (85.8) | 197 (94.7) | 366 (91.5) |
| Direct patient care |  |  |  |  |
| No | 53 (81.5) | 107 (84.3) | 200 (96.2) | 360 (90.0) |
| Yes | 12 (18.5) | 20 (15.8) | 8 (3.9) | 40 (10.0) |
| Direct reports |  |  |  |  |
| No | 43 (66.2) | 80 (63.0) | 141 (67.8) | 264 (66.0) |
| Yes | 22 (33.9) | 47 (37.0) | 67 (32.2) | 136 (34.0) |
| Telework pre-pandemic |  |  |  |  |
| No | 40 (61.5) | 69 (54.3) | 108 (51.9) | 217 (54.3) |
| Yes | 25 (38.5) | 58 (45.7) | 100 (48.1) | 183 (45.8) |
| Years worked at the organization |  |  |  |  |
| Less than 3 years | 17 (26.2) | 34 (26.8) | 63 (30.3) | 114 (28.5) |
| 3 years or more | 48 (73.9) | 93 (73.2) | 145 (69.7) | 286 (71.5) |
| Ergonomic supports |  |  |  |  |
| Less than 5 | 26 (40.0) | 44 (34.7) | 42 (20.2) | 112 (28.0) |
| All five | 39 (60.0) | 83 (65.4) | 166 (79.8) | 288 (72.0) |
| Weekly teleconferencing |  |  |  |  |
| Less than 2 hours | 35 (53.9) | 45 (35.5) | 71 (34.2) | 151 (37.58) |
| 2 or more hours | 30 (46.1) | 82 (64.5) | 137 (65.9) | 249 (62.3) |
| Work life balance [(mean (SD))] | 3.4 (0.8) | 3.7 (0.8) | 4.1 (0.7) | 3.8 (0.8) |
| Productivity score [(mean (SD))] | 3.4 (0.6) | 4.1 (0.6) | 4.6 (0.5) | 4.3 (0.7) |
| Organizational trust [(mean (SD))] | 3.6 (0.7) | 3.8 (0.7) | 4.0 (0.7) | 3.9 (0.7) |
| Flexibility score [(mean (SD))] | 3.7 (0.7) | 3.8 (0.8) | 3.8 (0.7) | 3.8 (0.7) |

<sup>a</sup> n=65 (16.3%) includes 60 (15.0%) respondents who preferred to telework less than half the time and 5 (1.3%) respondents who preferred to not telework at all post-pandemic.

For occupational characteristics, 32.0% of respondents were in a union or employee group with collective group representation, 91.5% had a full-time job status, 10.0% provided direct patient care, 34.0% had one or more direct reports, 45.8% had already teleworked at least partially immediately prior to the pandemic and 71.5% had worked at the organization for three years or more.

For e-working conditions, the majority of the study sample reported yes for all five ergonomic supports (72.0%) and two or more hours of teleconferencing per week (62.3%) (Table 2). For the e-work life scale, the mean score for the productivity domain (4.3 out of 5) was higher than the mean scores for the work-life balance (3.8), organisational trust (3.9), and flexibility domains (3.8).

Over half of all respondents reported that they would like to telework post-pandemic all the time (52.0%), while 31.8% reported over half but not all the time and 16.3% reported less than half the time or not at all (Table 2).

In the unadjusted models, using less than half the time or not at all as the reference group, having at least one other person over five years of age in the home during working hours had a small significant association with a higher preference telework over half but not all the time (OR = 1.91, 95% CI: 1.04 to 3.50) (Table 3). Being a provider of direct patient care was strongly associated with a lower preference to telework all the time (OR=0.18, 95% CI: 0.07 to 0.45). Having all five ergonomic supports compared to less than five was moderately associated with a higher preference to telework all of the time (OR=2.63, 95% CI: 1.45 to 4.80). Two or more hours of teleconferencing per week compared to less than two hours moderately associated with a higher preference to telework over half but not all the time (OR=2.13, 95% CI: 1.16 to 3.91) and all the time (OR=2.25, 95% CI: 1.28 to 3.96). For the productivity domain, a higher mean score was strongly associated with a higher preference to telework over half but not all the time (OR=5.21, 95% CI: 2.90 to 9.37), and even more strongly associated with a higher preference to telework all the time (OR=19.36, 95% CI: 9.94 to 37.71). The work-life balance (over half but not all the time: OR=1.09, 95% CI: 0.69 to 1.71; all the time: OR=1.59, 95% CI: 0.97 to 2.59) and organizational trust domains (over half but not all the time: OR=1.33, 95% CI: 0.81 to 2.19; all the time: OR=1.53, 95% CI: 0.91 to 2.59) had small non-significant associations with the outcome. No relationship was identified between the flexibility domain and the outcome (over half but not all the time: OR=1.01, 95% CI: 0.60 to 1.70; all the time: OR=1.07, 95% CI: 0.62 to 1.85).

**Table 3:** Association of sociodemographic and occupational characteristics and e-working conditions with telework preference, unadjusted and three step hierarchical adjusted multinomial logistic regression models (reference = less than half the time or not at all)

|  | Unadjusted Models |  | Adjusted models |  |
| --- | --- | --- | --- | --- |
|  | Over half but not all the time<br>OR (95% CI) | All the time<br>OR (95% CI) | Over half but not all the time<br>OR (95% CI) | All the time<br>OR (95% CI) |
| **Step 1: sociodemographic<sup>a</sup>** |  |  |  |  |
| Age group |  |  |  |  |
| 35 or less | 1.00 | 1.00 | 1.00 | 1.00 |
| 36 to 45 | 1.24 (0.56, 2.76) | 1.21 (0.57, 2.55) | 1.25 (0.54, 2.86) | 1.15 (0.53, 2.49) |
| 46 to 55 | 0.85 (0.38, 1.89) | 1.09 (0.52, 2.26) | 0.79 (0.35, 1.81) | 1.01 (0.48, 2.14) |
| 56 or more | 1.33 (0.51, 3.50) | 1.02 (0.40, 2.57) | 1.21 (0.45, 3.24) | 0.94 (0.37, 2.39) |
| Gender identity |  |  |  |  |
| Woman or non-binary | 1.00 | 1.00 | 1.00 | 1.00 |
| Man | 0.64 (0.28, 1.44) | 0.99 (0.48, 2.03) | 0.53 (0.23, 1.24) | 0.92 (0.44, 1.92) |
| Children under 5 years of age |  |  |  |  |
| None | 1.00 | 1.00 | 1.00 | 1.00 |
| One or more | 1.49 (0.71, 3.13) | 1.36 (0.67, 2.75) | 1.62 (0.74, 3.52) | 1.36 (0.66, 2.83) |
| People over 5 years of age |  |  |  |  |
| None | 1.00 | 1.00 | 1.00 | 1.00 |
| One or more | 1.91 (1.04, 3.50) | 1.29 (0.74, 2.26) | 2.07 (1.11, 3.87) | 1.36 (0.77, 2.42) |
| Disability or long term condition |  |  |  |  |
| No | 1.00 | 1.00 | 1.00 | 1.00 |
| Yes | 1.03 (0.39, 2.68) | 1.56 (0.66, 3.72) | 1.06 (0.40, 2.82) | 1.63 (0.68, 3.92) |
| **Step 2: occupational<sup>b</sup>** |  |  |  |  |
| Union |  |  |  |  |
| No | 1.00 | 1.00 | 1.00 | 1.00 |
| Yes | 1.07 (0.57, 2.01) | 0.81 (0.45, 1.47) | 1.26 (0.57, 2.77) | 1.43 (0.69, 2.98) |
| Job status |  |  |  |  |
| Not full time | 1.00 | 1.00 | 1.00 | 1.00 |
| Full time | 0.50 (0.18, 1.43) | 1.49 (0.50, 4.47) | 0.47 (0.15, 1.48) | 1.22 (0.36, 4.10) |
| Direct patient care |  |  |  |  |
| No | 1.00 | 1.00 | 1.00 | 1.00 |
| Yes | 0.83 (0.38, 1.81) | 0.18 (0.07, 0.45) | 0.65 (0.25, 1.71) | 0.14 (0.05, 0.40) |
| Direct reports |  |  |  |  |
| No | 1.00 | 1.00 | 1.00 | 1.00 |
| Yes | 1.15 (0.61, 2.15) | 0.93 (0.51, 1.68) | 1.04 (0.53, 2.01) | 0.79 (0.42, 1.48) |
| Telework pre-pandemic |  |  |  |  |
| No | 1.00 | 1.00 | 1.00 | 1.00 |
| Yes | 1.34 (0.73, 2.47) | 1.48 (0.84, 2.62) | 1.41 (0.73, 2.74) | 1.42 (0.77, 2.64) |
| Years worked at the organization |  |  |  |  |
| Less than 3 years | 1.00 | 1.00 | 1.00 | 1.00 |
| 3 years or more | 0.97 (0.49, 1.91) | 0.82 (0.44, 1.53) | 0.81 (0.36, 1.79) | 0.80 (0.38, 1.67) |
| **Step 3: e-working conditions<sup>c</sup>** |  |  |  |  |
| Ergonomic supports |  |  |  |  |
| Less than 5 | 1.00 | 1.00 | 1.00 | 1.00 |
| All five | 1.26 (0.68, 2.33) | 2.63 (1.45, 4.80) | 0.54 (0.23, 1.26) | 0.66 (0.26, 1.66) |
| Weekly teleconferencing |  |  |  |  |
| Less than 2 hours | 1.00 | 1.00 | 1.00 | 1.00 |
| 2 or more hours | 2.13 (1.16, 3.91) | 2.25 (1.28, 3.96) | 2.85 (1.25 to 6.46) | 3.69 (1.52, 8.92) |
| Work life balance | 1.09 (0.69, 1.71) | 1.59 (0.97, 2.59) | 1.52 (0.87, 2.66) | 3.06 (1.65, 5.65) |
| Productivity | 5.21 (2.90, 9.37) | 19.36 (9.94, 37.71) | 7.46 (3.70, 15.05) | 28.84 (13.22, 62.92) |
| Organizational trust | 1.33 (0.81, 2.19) | 1.53 (0.91, 2.59) | 1.54 (0.89, 2.69) | 1.81 (0.99, 3.30) |
| Flexibility score | 1.01 (0.60, 1.70) | 1.07 (0.62, 1.85) | 0.73 (0.38, 1.41) | 0.61 (0.31, 1.22) |

<sup>a</sup>The step 1 adjusted model includes all sociodemographic characteristics. AIC = 817.09.
<sup>b</sup>The step 2 adjusted model includes all sociodemographic and occupational characteristics. AIC = 811.93.
<sup>c</sup>The step 3 adjusted model includes all sociodemographic characteristics, occupational characteristics and e-working conditions. AIC = 637.40

After adjusting for other sociodemographic factors (step 1 adjusted model, Table 3), the relationship of each sociodemographic factor with telework preference was similar to the unadjusted results. Having at least one other person over five years of age in the home during working hours remained moderately associated with a higher preference to telework over half but not all the time (OR=2.07, 95% CI: 1.11 to 3.87).

After adjusting for sociodemographic and other occupational factors (step 2 adjusted model), the relationship of each occupational factor with telework preference was also similar to the unadjusted results. Being a provider of direct patient care remained strongly associated with a lower preference to telework all the time (OR=0.14, 95% CI: 0.05 to 0.40).

After adjusting for sociodemographic, occupational factors, and other e-working conditions (step 3 adjusted model), ergonomic supports were no longer significantly associated with telework preference (over half but not all the time: OR=0.54, 95% CI: 0.23 to 1.26; all the time: OR=0.66, 95% CI: 0.26 to 1.66). Effect estimates for the weekly teleconferencing, work-life balance, productivity, and organizational trust variables were similar but stronger (i.e. further away from ‘1’) compared to the unadjusted results. Two or more hours of teleconferencing per week compared to less than two hours was moderately associated with a higher preference to telework over half but not all the time (OR=2.85, 95% CI: 1.25 to 6.46) and all the time (OR=3.69, 95% CI: 1.52 to 8.92). Higher work-life balance was moderately associated with a higher preference to telework all the time (OR = 3.06, 95% CI: 1.65, 5.65). Higher productivity was strongly associated with a higher preference to telework more than half but less than all the time (OR = 7.46, 95% CI: 3.70 to 15.05) and even more strongly associated with a preference to telework all the time (OR = 28.84, 95% CI: 13.22 to 62.92).

The AIC value decreased across each successive step of the adjusted hierarchical models indicating that the model fit improved with each step.

## Discussion

Employees’ telework preference for the post-pandemic period is a pressing issue for society and employers. Ensuring fit between employee need and the work arrangement has implications for employees and organizations including employee health, job performance, recruitment and retention. 19,20 This study sought to expand current knowledge in this area on prevalence and determinants in the healthcare sector for the post-pandemic context, as well as by using a hierarchical multiple regression approach to prevent biases that can occur due to entering both proximal and distal determinants into the same model at once. A key finding was that 99% of the study sample of healthcare employees who teleworked at some point during the pandemic preferred to continue teleworking for either all or some of the time post-pandemic. This estimate is higher than the estimate of 90% reported by an Australian survey of corporate and non-clinical employees of a large local health district who adapted home-based telework during the pandemic. This difference may be due to the inclusion of non-home based telework including virtual work performed in a clinic or hospital in this study’s telework preference measure. It is also higher than estimates from other multi-sector studies where the proportion of employees preferring to telework post-pandemic ranged from 91% for Canadians who newly teleworked during the pandemic and 39% to 82% for non-Canadian working populations. 3,5–11 This may be due to a higher proportion of jobs with a capacity for telework under normal circumstances, a higher average income, and higher job security, and longer commute times in the current study sample; as well as the inclusion of non-home based telework in the telework preference measure as mentioned earlier. It is notable that 45.8% of the current study sample teleworked at least partially immediately prior to the pandemic – indicating a high capacity for telework arrangements under normal circumstances. Higher income may be associated with an increased preference to telework due to the ability to afford better telework conditions such as a larger living space or a more comfortable work station. Higher job security may be associated with an increased preference to telework; as higher job security may lessen concerns of experiencing negative occupational outcomes like lack of career progression due to being a teleworker. 13 Lastly, a longer commute has been associated with a preference to telework. 12 As the employer in the current study covers a large geographical service area, longer commuting times are likely common among its employees.

Another key finding of the current study is that there was a relatively even split between employees who prefer to telework all the time (52.0%) and employees who prefer a hybrid arrangement with both on-site and telework components post-pandemic, with teleworking over half the time but not all the time (31.8%) being a more popular arrangement than teleworking less than half the time (15.0%). This finding is similar to a multi-sector American study that found that of employees who want to telework post-pandemic, preferences are split between wanting to telework all of the time (50%) and some but not all the time (50%). 5 However, it varies from a multi-sector study of Canadian employees, where employees were five times more likely to prefer a post-pandemic hybrid tele-work/on-site work arrangement over an exclusively teleworking arrangement. 11 It also varies from multi-sector studies of Hong Kong and South African employees where hybrid post-pandemic telework schedules ranging from one to three days a week of teleworking were more popular than exclusively teleworking arrangements. 7,8,10 Reasons for a majority preference to telework for all hours in the current study sample, may be due to the same mechanisms discussed above: higher telework capacity under normal circumstances, higher income and job security, and longer commutes, as well as the inclusion of non-home based telework in the telework preference measure.

In this study, strong determinants of telework preference included being a provider of direct patient care and productivity while teleworking. Moderate determinants included number of weekly teleconference hours, work-life balance and having one or more people over five years of age in the home while teleworking.

Being a provider of direct patient care was associated with a lower preference to telework all of the time compared to less than half of the time or not at all. This finding may be due to the limitations of virtual settings for healthcare delivery including difficulty reading body language or social cues 28, making accurate diagnoses, and delivering treatments especially those that involve physical procedures. Of importance to note, is that the vast majority of direct patient care providers preferred a hybrid work arrangement with the most common preference being over half the time but not all the time spent teleworking followed by some of the time but less than half the time. This finding is in compatible with the results of a recent integrative review that found that overall, healthcare providers were satisfied with the use of telehealth during the pandemic and willing to continue using telehealth following the pandemic to provide direct patient care, particularly for follow up visits. 29

Employees who scored higher on work-life balance and/or productivity while teleworking, were more likely to prefer a future work arrangement with a greater number of hours spent teleworking. The current finding for productivity was replicated in a recent multi-sector survey of Canadians who teleworked for most of their work hours in the first year of the pandemic. 11 Both work-life balance and productivity while teleworking may influence post-pandemic telework preference via job satisfaction, general health and well being and mental health. 23,30 In addition, employees who scored high on work-life balance or productivity while teleworking, may anticipate that returning to on-site work post-pandemic will negatively impact their work-life balance or productivity, leading them to strongly prefer to continue teleworking for most or all of their work hours.

Two or more hours of weekly teleconference activity was associated with a preference to telework most or all the time compared to less than half the time. Employees with less than two hours of weekly teleconference activity may be more likely to feel socially isolated from their work colleagues and this may in turn increase preference to partially return to on site work with a greater number of hours on site. Alternatively, spending over two hours a week on teleconference activities may be reflective of effective adaptation of virtual teamwork or telehealth practices which in turn may be associated with same or increased productivity and an increased preference to continue teleworking for the majority or all of the time.

Having one or more people over the age of five in the home during work hours was associated with a preference to telework over half the time but not all the time compared to less than half the time or not at all. Employees with no other people over the age of five in the home during working hours may experience more social isolation while teleworking and this may increase preference to partially return to on site work with a greater number of hours on site.

Having one or more people under the age of five in the home during working hours had a small non-significant association with a preference to telework for most or all of the time over less than half the time or not at all. Other studies of Vietnamese and American employees point to having young children in the home as a positive determinant of having a preference or willingness to telework post-pandemic, especially among women. 12,31 The current findings do not contradict this, but having young children in the home may have been a smaller determinant in the current study sample, especially compared to occupational factors and e-working conditions, due to a more equitable balance in the distribution of unpaid labour across spousal partners with young children in the current study sample of Canadian healthcare employees compared to Vietnamese employees. 32,33. Another possible reason is that due to Canada’s more generous parental leave benefits in comparison to the United States, Canadian employees with an infant in the home, may be more likely to have one or more parents on parental leave than their American counterparts. This may lessen the need for home-based telework as a means to provide infant childcare or related unpaid working activities during working hours or time that would otherwise be spent commuting.

In the current study, there was no significant association between gender identity and telework preference. A recent Canadian multi-sector survey had similar findings 4 while Malaysian and American multi-sector surveys found that women are more likely to prefer to telework or to prefer to telework for a greater proportion of their work hours post-pandemic than men. 12,31 The lack of a strong effect for gender identify in the current study may be due to a more equitable distribution of unpaid labour across spousal partners in the current study of healthcare employees in British Columbia Canada compared to studies conducted in other countries. 32 Alternatively, it may be due to a lower concern among Canadian men regarding potential teleworking disadvantages compared to men in other countries. Research on Lithuanian employees conducted during the pandemic found that men were less accepting of role-conflict such as work disturbance by family members during home-based telework, and that men were more concerned about improper assessment of their competency or performance by their supervisors and constraints on career opportunities due to telework. 34 These disadvantages may have been of lower concern to men in the current study sample due to higher job security or work and home environments that are comparatively less patriarchal. 35 This latter mechanism may be particularly prominent in female dominant sectors such as healthcare.

A survey broadly aimed at Portuguese teleworkers during the first pandemic wave used the e-work life scale flexibility and organizational trust subscales to examine determinants of job satisfaction. 36 Higher organizational trust was a major predictor and higher work flexibility was a good predictor of higher telework satisfaction. 36 In the current study, neither of these subscales were moderate or strong determinants of telework preference. Discrepancies in these findings may be due to differences in the constructs of telework satisfaction and telework preference or differences in the study samples. Experiences of organizational trust and work flexibility may vary more greatly across employees from different organizations and sectors as in the Portuguese study than among employees of the single healthcare organization examined here. Further, the flexibility and organizational trust scores were higher in the current study and this may have resulted in a ceiling effect for these variables.

Two major strengths of this study include: 1) the consideration of potential sociodemographic, occupational and e-working determinants of healthcare workers’ post-pandemic telework preference and 2) a hierarchical modelling approach to prevent bias in effect estimates for distal determinants due to adjustment for more proximal determinants in the same model. However, there are some limitations to consider. First, while a participation rate of 36.4% was achieved, information on non-respondents necessary to determine the representativeness of the study sample was not available for analysis. Second, no survey questions addressed commuting practices when working on site. Long commutes may have explained the high preference to telework most or all of the time in the study sample. Third, the target population included employees of the large regional health authority who teleworked during the pandemic.

The results are most generalizable to healthcare and other large organizations in high income countries, particularly those with a workforce or service area distributed across multiple communities, and a highly educated or skilled workforce with job tasks that can be performed remotely using information technology. Lastly, employees’ telework preference may evolve over time. It is important to contextualize the findings relative to the data collection period which was during the latter part of the third pandemic wave.

These findings have multiple considerations for policy and workforce planning. In some organizations, a majority of employees want to continue teleworking for most or all of their work hours, even after risk of COVID-19 transmission in the work place is significantly reduced. This has implications for on-site space allocation, support equipment and services for telework, work-home boundary management, networking facilitation and productivity management for telework arrangements, and return to work planning particularly the allowance of continued telework arrangements for employee satisfaction and retention. The findings indicate a special consideration of continuation of telework for providers of direct patient care; and that hybrid work arrangements with more onsite work hours but not all the time may be more popular among these employees than among employees who do not provide direct patient care. With an increasing shift towards virtual health care delivery, this finding still emphasizes the importance of in-person direct health care delivery. However, the hybrid arrangement might enable direct patient care providers to perform a subset of tasks at home or virtually. Another consideration is that long-term continuation of telework practices established during the pandemic may have implications for the onsite work environment as well as employees unable to perform their job via telework, which is common in healthcare. These include changes to management and collaboration practices and a potential lack of onsite presence by senior employees or decision makers as well as changes to moral or satisfaction with the work arrangement or overall job for exclusively on site employees. Lastly, work arrangements with at least some on-site components may be particularly important for employees who experience low productivity, low work-life balance, or infrequent or ineffective virtual communication or socialization with colleagues while teleworking. Alternatively, addressing productivity, work-life balance, or virtual communication or socialization issues that arise while teleworking, may help to increase employees’ preference to continue teleworking post-pandemic.

## Data Availability

In accordance with the ethics agreement and to protect participant anonymity, data is not available for sharing.

